# A Computer Simulation Study on novel Corona Virus Transmission among the People in a Queue

**DOI:** 10.1101/2020.05.16.20104489

**Authors:** Santhosh Samuel Mathews

## Abstract

The World Health Organization (WHO) on March 11, 2020, has declared the novel Corona virus (COVID-19) outbreak a global pandemic. It is essential to understand how coronavirus transmits from one person to another and this knowledge will help protect the vulnerable and limit the spread of the Corona virus. The mode of respiratory transmission of Corona virus is not completely understood as of date. Using a computer simulation, this paper analyses the probability of spreading of Corona virus through air among the people who are standing in a queue. The parameters such as the diameter of the virus particle, room temperature, relative humidity, height of the person, distance between the people and the waiting time in the queue are considered in the computer model to determine the distribution of Corona virus and hence identify the risk factor of spreading the Covid-19. This paper describes the possibilities of getting infectious when a Covid-19 infected person present in a queue and the impact on the waiting time, distance between people and the position in the queue on the transmission of Corona virus.

## Introduction

Covid-19 is a human to human spreading disease due to corona virus and experiences cough, fever with high body temperature and difficulty in breathing and the serious cases can lead to pneumonia, kidney failure or even death. The World Health Organization (WHO) on March 11, 2020, has declared the novel Corona virus (COVID-19) outbreak a global pandemic. Most respiratory viruses are most contagious when a person has symptoms. However, there is growing evidence to suggest that the virus might also spread during the incubation period, before a person develops any symptoms.

As the Human Corona virus (HCoV) is new and not much studies has been performed, all possibilities of transmission of corona virus corresponding to Covid-19 is not completely known. Due to the high spreading rate of Covid-19 requires more attention to understand all transmission methods of Corona virus. This paper analyses the possibility of transmission of Corona virus through air especially the chances of spreading when people who are standing in the queue.

### Transmission

All mechanisms of human oro-nasal activity such as breathing, talking, laughing, coughing and sneezing produce particles within the inhalable range for humans of <1 to >100 μm.[2] Droplets are exhaled when we breathe, speak, laugh, cough, or sneeze, and the size of the droplet determines whether it is considered "droplet” or "aerosol”. Droplets >5 μm in diameter are generally referred to as droplets, and those ≤5 μm are generally referred to as aerosols. The droplet size distinction is important; the smaller the droplet, the longer and farther it may travel in the air and the more likely it can avoid becoming trapped by mucus or cilia and reach the alveoli of the lower respiratory tract.[3]

One of the recent studies in China have shown that the air transmission of Human Corona virus via droplets in air-conditioned rooms are possible.[24] The common respiratory influenza virus and respiratory syncytial virus are considered to be spread mostly by droplets; however, aerosol transmission also is believed to occur. Respiratory spread of virus during the outbreak of severe acute respiratory syndrome (SARS) in China in 2003 is thought to have principally occurred via respiratory droplets. However, aerosol spread of this virus has also been postulated, and this virus has been shown to occur in laboratory-generated aerosols.[3]

The respiratory droplet concentration depends on medical parameters (breathing capacity, lungs capacity) which are depend on human height, weight, gender, age etc. The spread probability of respiratory disease depend on the droplet characteristics, which includes droplet size, concentration, droplet velocity, temperature, type of pathogen, activity of pathogen etc. It has been shown that droplet pathogens do not exclusively airborne or droplet-borne transmission but both methods simultaneously. The size of breathing droplets from healthy people is 0.1 to 8uM and the patient is 0.05 to 10uM.[4] The mode of respiratory transmission of SARS-CoV-2 is not completely understood.[3]

### Size and shape

Scientists have already used electron microscopes to measure how big the corona virus is. Coronavirus virions (or ‘particles’) are spherical particles with diameters of approximately 125 nm (0.125 microns). The smallest particles are 0.06 microns, and the largest are 0.14 microns. [5-6] For the comparison with influenza virus, Influenza type A viruses are very similar in structure to influenza viruses types B, C, and D. The Influenza virus particle (also called the virion) is 80–120 nanometres in diameter such that the smallest virions adopt an elliptical shape.[1]

### Effect of temperature and Humidity on enveloped viruses

Corona viruses are enveloped viruses with a positive sense, single stranded RNA genome. Influenza viruses are also enveloped viruses.[7] In some studies observed that the Human coronavirus, pseudorabies virus, and rotavirus are most stable at intermediate RH. The first two are enveloped, while mature rotaviruses are usually non-enveloped. [9] In these studies, the decay of virus infectivity increased rapidly at relative humidity >40%. The increased survival of influenza virus in aerosols at low relative humidity has been suggested as a factor that accounts for the seasonality of influenza.[10]

Three mechanisms could potentially explain the observed influence of RH on transmission. The first acts at the level of the host: breathing dry air could cause desiccation of the nasal mucosa, leading to epithelial damage and/or reduced mucociliary clearance, which would in turn render the host more susceptible to respiratory virus infections. Long-term exposure to dry air is likely to affect influenza virus growth in the upper respiratory tract, and may indeed play a role in influenza seasonality.

The second mechanism acts at the level of the virus particle. The stability of influenza virions in an aerosol has been reported to vary with RH. The most recent of these reports [12] shows viral stability to be maximal at low RH (20%–40%), minimal at intermediate RH (50%), and high at elevated RH (60%–80%). A detailed study conducted by Anice C.L, shows the stability of the virus in aerosols is a key determinant of influenza virus transmission (with the exception of the absence of transmission at high RH).

The third mechanism acts at the level of the vehicle, the respiratory droplet. At low RH, evaporation of water from exhaled bioaerosols would occur rapidly, leading to the formation of droplet nuclei; conversely, at high RH, small respiratory droplets would take on water, increase in size and settle more quickly out of the air [11].

There are a few studies conducted to understand the effect of ambient temperature on the transmission of enveloped viruses such as influenza A and influenza B. The temperatures less than 20 degrees shows increased transmission efficiency of enveloped viruses explains the reason for seasonal flue. [8][11] The author of this paper hypothesis that this is primarily due to the evaporation of water contents from the envelop at higher temperature and the virus will be inactive. The term "critical diameter” is defined as the smallest diameter of the enveloped virus including its water envelop at which the virus stay active. At higher ambient temperatures, the virus envelop evaporate slowly and when it reaches the critical diameter it become inactive.

### Viruses in Human exhale

As of today, no data is available about the number of virus particles that can be exhaled by a Covid-19 patient. One experiment conducted by David A et.al found that some normal human subjects expire many more bioaerosol particles than other individuals during quiet breathing and therefore bear the burden of production of exhaled bioaerosols. [22] The amount of virus particles expelled by an individual depends on their anatomy such as lungs capacity, lung airway surface properties etc. In this computer simulation, the amount virus particles exhaled is variable and is defined in one of the model parameters.

### Risk of infection

The amount of virus necessary to make a person sick is called the infectious dose. Viruses with low infectious doses are especially contagious in populations without significant immunity. The minimum infectious dose of SARS-CoV-2, the virus that causes Covid-19, is unknown so far, but researchers suspect it is low.

A high infectious dose may lead to a higher viral load, which can impact the severity of Covid-19 symptoms. Depending on the virus, people need to be exposed to as little as 10 virus particles — for example, for influenza viruses — or as many as thousands for other human viruses to get infected.[14][15]

## Materials and methods

The computer model is written in Spyder IDE with Python programming language. The Python libraries are extensively used for this computer model.

### Parameters for modelling the airborne spread of Corona virus

#### Excretion of corona virus

At rest about 0.5L of tidal air is inhaled and exhaled with each breath. During heavy work, the tidal volume may exceed three times this amount. Resting adult breathes about 12 times a minute. In the computer model, the breath rate is denoted as Breath_rate_ and set the default value as 12 per minute. ie. Exhale once in 5 seconds. As of date, more details of Corona virus is not known to us, one of the closest respiratory disease Influenza is taken as the reference in this computer simulation. Concentrations in exhaled breath samples ranged from 48 to 300 influenza virus RNA copies per filter on the positive samples, corresponding to exhaled breath generation rates ranging from 3.2 to 20 influenza virus RNA copies per minute.[13] The same set of values are assigned to the model parameter V_exhale_ for the simulation of virus exhale. In operation, the simulation model will excrete V_exhale_ number of virus particles in every exhale whose rate is determined by Breath_rate_. As a default value, the number of particles exhaled by the infected person is set to 5 particles per exhale.

#### Waiting time in the queue

The waiting time in the queue is an import model parameter which decides the number of virus particles expelled to the surrounding by the infected person as well as the number of particles inhaled by the health person from the surrounding. The model parameter Q_Wait_ defines the waiting time of the infected person in the queue (which is same as the waiting time of healthy people in the same queue) in seconds.

#### Distance between people in the queue

The virus particles excreted by the infected person in the queue will be in the surrounding air or settled on the ground. One of the factors that decides the probability of inhaling the ejected particles suspended in the surrounding area by a healthy person is the distance from the infected person in the queue. The Q_Dist_ model parameter determines the distance between the people in the queue.

#### Consume factor

A healthy person in the queue is vulnerable to the virus particles that suspended in the air. The probability of getting the disease is based on the number of virus particles inhaled which is above the infectious dose of Covid-19. The model parameter Cons_fact_ defines the fraction of the total number of particles that inhaled by the person in the queue. The value of Cons_fact_ is always less than or equal to 1. The consume factor is used to determine the amount of viruses inhaled by the healthy person standing in the queue and remove the exact amount from the environment.

#### Scattering factor

The computer model assumes that the suspended particles in the air will free fall under the influence of the gravitational force through the air column. However, the movement of people and objects in the surrounding may scatter the virus particles and deviate them from their trajectory. The model parameter Scatter_fact_ is defined as the fraction of the total number of virus particles lost when they travel at a distance of 0.1 meter. The Scatter_fact_ is always less than or equal to 1.

#### Height of the person

The virus particles exhaled out or inhaled in through the nose or mouth of the person during breathing, talking, coughing or sneezing. So the viruses exhaled or inhaled is from the area of air at their height. The height of the infected person PatientHt and that of the healthy person HealthyHt are defined as model parameters.

#### Creating virus in the simulation

In real life, the virus particle can be either aerosol or droplet based on the way it is released from the infected person. The physical characteristics of the particle may vary from aerosol to droplet but the computer model will consider the diameter of the particle as its main parameter. The virus particle is created with the given diameter in the computer simulation. The diameter can be in random which will be in between in VirusDia_Start_ and VirusDia_End_ where the smaller diameter particles will behave as aerosol and larger particles as droplets. The number of viruses created per exhale is determined by the V_exhale_ while the position of release in the x axis of these particles are determined by the position of the infected person in the queue. Similarly the height of release of particle is determined by the height of the infected person as it is released during the talk, cough or sneeze or even in tidal breathing.

#### Settling velocity

Due to the gravitational force, the excreted virus particles will free fall to the ground and the time taken to settle them is decided by the setting velocity of the particle. Settling velocity of a droplet or an airborne virus is the velocity reached by it as it falls through air and dependent on its size. The settling velocity can be determined by the equation [16]

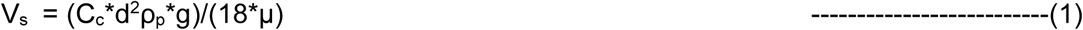

Where V_s_ is the settling velocity

C_c_ = Cunningham (slip) correction factor
d = Particle diameter in meter
ρ_p_ = density of particle, unit density = 1 kg/m^3^
g = acceleration of gravity, 9.81 m/s^-2^
μ = viscosity of gas, air = 1.81e-05 kg/m·s

As of date, the density of Corona virus is not known and has taken the same as that of influenza virus. Density of influenza virus is 1.1 g/cm^3^. [17].

Cunningham slip factor can be written as [16]

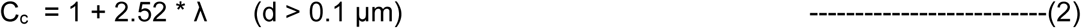

Where λ is the mean free path of molecules in the fluid. For air it is 0.067uM.

#### Survival of corona virus

There are some environmental parameters that determines the survival of air suspended virus particles which mainly includes humidity and temperature.

#### Humidity

As of today, no data is available about the relation of ambient Relative Humidity with the survival of Corona virus and hence taken the details of Influenza virus mentioned in some of the studies. The viability of infectious influenza viruses decreases over time and is affected by environmental variables such as temperature, humidity, and UV radiation. The inactivation rate (k) per minute, derived from experimental data on airborne Influenza A virus, is linearly correlated with RH, following the relationship

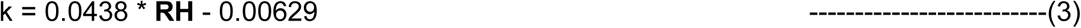

with an r2 of 0.977 and p-values for the model, intercept, and slope of 0.0015, 0.059, and 0.0015, respectively.[8][18] The ambient relative humidity in percentage is defined in the model parameter Room_RH_.

#### Temperature

The exact mechanism of the survival of enveloped virus at higher ambient temperature is not known fully today. In the case of droplets with the larger diameter, the higher temperature causes the evaporation of the virus envelop that will reduce the diameter and hence allow them to suspend in the air for more time. At the same time, in the case of droplets with smaller diameter causes the removal of water contents in the envelop due to higher temperature and hence the virus decay is due to changes in droplet chemistry due to the evaporation. A full mechanistic explanation may be revealed by future research examining the chemistry of evaporating droplets and the physiological effect on virus.[20] It is important to note, however, that a very small seasonal change in transmission rate is thought to be sufficient to drive large changes in influenza incidence, due to amplification through dynamical resonance. Thus, relatively brief exposure, or extended exposure of relatively few individuals, to cold temperatures may have a large impact on viral circulation. [19] Since the Corona virus is also an enveloped virus like Influenza virus, hence it is important to consider both relative humidity and room temperature in this computer simulation.

The ambient temperature is represented as Room_Temp_ which is in degree centigrade in the computer model. This computer model uses Python pyvap module to calculate the rate of evaporation of water contents in the Corona virus.

### Risk of infection of the person in the queue

The amount of virus necessary to make a person sick is called the infectious dose. Viruses with low infectious doses are especially contagious in populations without significant immunity. The minimum infectious dose of SARS-CoV-2, the virus that causes Covid-19, is unknown so far, but researchers suspect it is low.[21] Since the infectious dose of Covid-19 is not known as of date, the model parameter **InfectiousDose** is not used in the current study.

## Computer simulation

The viruses accumulated due to exhale of the infected person

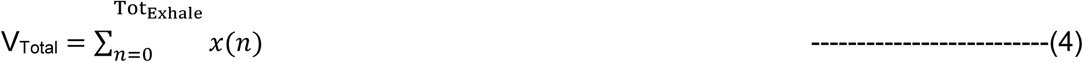

Where V_Total_ is the total viruses exhaled after Tot_Exhale_ number of exhales and *x*(*n*) is the number of viruses exhaled by the infected person at the n^th^ exhale. The time taken per exhale in seconds K can be calculated as

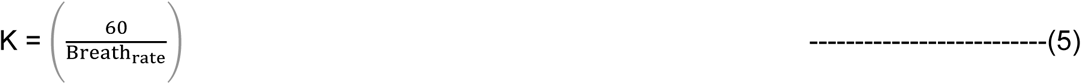

where Breath_rate_ is the breath rate in minutes. In this computer model, the number of viruses excreted per exhale *x*(*n*) remains same and is defined in the model parameter V_exhale_. The person in a queue remains in the same position for a duration of Q_Wait_ seconds and excrete the virus to the same air column during this period if he or she is infected by Corona virus. The number of exhale during this period is

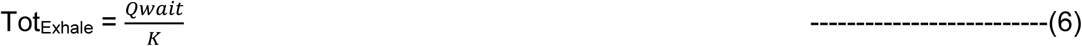

where K can be calculated using the equation (5). From equations (4) and (6), the total viruses excreted in a particular air columns can be re-written as

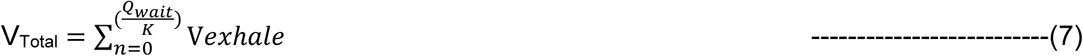

Each person in a queue wait at the same position during Q_wait_, then shift to the next position which causes of leaving the contaminated area of air column behind when the infected person advances one step in the queue. Mathematically this can be written as

Pos(t) = x(t) where x(t) is the position of the infected person in the queue at the time t t_n_ = t_n−1_+Q_wait_ and 0 ≤ t_n_ ≤ T_sim_ where T_sim_ is the simulation time in the computer model which is same as the time taken by the person to move from the end of the queue to the beginning of the queue. Also, the position of the healthy person who is behind the infected person in the queue can be defined mathematically as

Pos(t) = y(t) where y(t) is the position of the healthy person at the time t in the queue Where t_n_ = t_n−1_ + Q_wait_ and

Q_wait_ ≤ t_n_ ≤ T_sim_

The position of the healthy people is also can be written with respect to the position of the infected person as

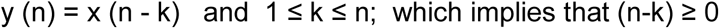

k represents the position of the people from the infected person, ie. the value of k=1 represents the healthy person just behind the infected person, k=2, second position from the infected person and so on.

The time taken by a person to reach the next position can be written as

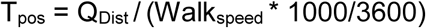

At any point of time, the number of viruses in a section will be decided by the distance travelled by a virus particle over the time T.

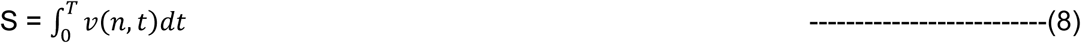

Where S is the distance travelled by the n^th^ particle in meters over a period of time T and *v*(*n, t*) is the setting velocity in ms^-1^ of the n^th^ particle at t^th^ time which can be calculated by the equation (1). Please note that as per the equation (1), the settling velocity is independent of time.

As the particles free fall, there is a possibility of scattering of some particles and this can be due to the movement of non-living or living organisms in the air column. The scattering factor Scatter_fact_ is defined as the factor of removed viruses when they travel 0.1 meter in the air.

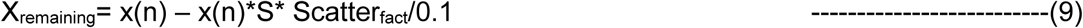

Where X_remaining_ is the remaining viruses in the cluster, *x*(*n*) is the number of particles in the n^th^ cluster and S is the distance travelled in meters and Scatter_fact_ is the scatter factor per 0.1 meter. S can be calculated from the equation (8).

In the case of still air, the velocity of the particle is only due to the influence of settling velocity (as per the equation (1)), the virus particles will be distributed across the vertical air column in front of the infected person. For the simulation purpose, the air column is divided into small sections of height 0.01 meters and the accumulated virus particles in a section is calculated using the formula,

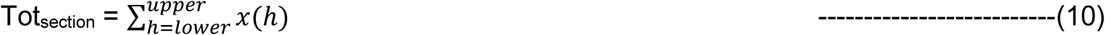

Where Tot_section_ is the total virus particles in a section

*lower* is the lower height of the section
*upper* is the upper height of the section which is same as *lower +* 0.01 meters

and

*x*(*h*) is the number of viruses at a height h.

When the particle height reaches to ***MinHeight*** of 0.25 meter or less from the ground, it is assumed that the virus is settled on the surface and henceforth the airborne transmission can be neglected. Therefore the equation (10) is re-written as

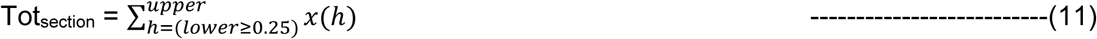

Where *lower* is greater than or equal to MinHeight.

The viruses inhaled by a healthy person when he or she is in the contaminated air column, can be written from the equation (10) as

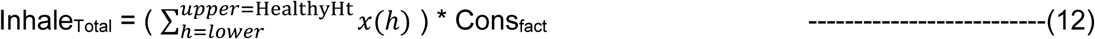

where *upper* will be the height of the person, *lower* is *upper* - 0.01 meters, Cons_fact_ is the consume factor which is defined as the factor of total viruses that will be inhaled by a person in every breath.

## Validation of the model

The results and observations from computer simulation are validated against the results of various experiments conducted by other researchers on the same or similar pandemic problems.

### Settling velocity

To validate the calculation of the settling velocity of the aerosol or the droplet, determined the same with the particles of size from 0.1 μM to 100 μM and found close to the values mentioned in other sources.

**Table 1:** Settling velocity calculated by the computer model

| Sl. no | Dia of the particle ($\mu\text{M}$) | Settling velocity calculated by the model (m/s) |
| --- | --- | --- |
| 1 | 0.1 | 8.3198e-07 |
| 2 | 0.5 | 1.0125e-05 |
| 3 | 1.0 | 3.538e-05 |
| 4 | 5.0 | 7.825e-04 |
| 5 | 10.0 | 3.0788e-03 |
| 6 | 20.0 | 1.2213e-02 |
| 7 | 30.0 | 2.7403e-02 |
| 8 | 50.0 | 7.595e-02 |
| 9 | 75.0 | 1.7069e-01 |
| 10 | 100.0 | 3.0328e-01 |

### RH and Temperature

The existence of virus particles with various temperature and relative humidity are determined by the computer model. The number of viruses inhaled by the person in the queue is determined with RH from 20% to 100% and with ambient temperatures 14° C, 19° C, 25° C and 37° C. The observations are plotted in the following graph.

**Figure 1:**
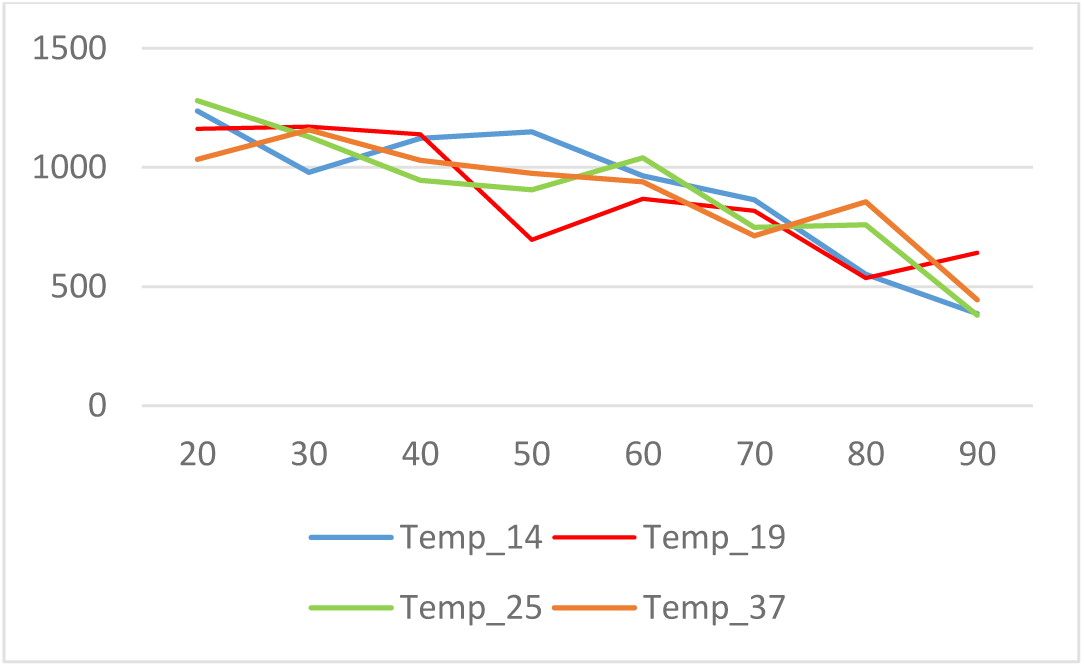
Relation of number inhaled virus with temperature and humidity.

Inhaled virus particles vs Relative Humidity (%)

X-axis: Relative Humidity (%) Y-axis: Number of particles inhaled

In this computer model, as the humidity increases, the number of inhaled viruses reduces especially above 70% of humidity. This is an expected result and is mentioned in some of the research papers.[8][12][23]. However, the temperatures from 14°C to 37°C are not having much effect. This can be explained by the fact that the evaporation of the enveloped water content due to the ambient temperature. The water content in the enveloped virus will evaporate based on the temperature but it will be a slow process. The slight increase in the number of viruses at higher RH (say 80%) and higher temperature is due to the evaporation of water contents from the droplets. This evaporation will cause the overall reduction in the diameter of enveloped viruses, reducing the settling velocity and hence allow them to suspend in the air for more time. Also it is worth to note that, the duration of operation of the model depends on the time taken by the infected person in the queue, ie. Moving from the end to the front of the queue including the waiting time in between. This time may not be sufficient to evaporate the entire water content of the virus and hence may not inactivate the virus at the given temperature.

### Assumptions

This study is concentrated on the possibilities of spreading of Corona viruses among the people who are standing in a queue especially in indoor controlled environment. More specifically, this deals with possibilities of spreading of Covid-19 when people are standing in the queue such as in front of the check-in counter in an airport, check-out counter in a shopping mall, buffet counter in a restaurant, ticket counter in a public transport station etc. Even though the computer model accommodates the effect on drift velocity of virus particles due to the cross wind and front wind, this study assumed that the virus particles are traversed and settled in still air and subsequently the cross wind and front wind velocity model parameters are set to zero in all comparative studies that mentioned in this paper.

The model considers the effect of environmental factors such as temperature and humidity and the effect can be studied by changing appropriate model parameters. However, in a controlled environment, the temperature and humidity are set for the comfort of the human by artificial means and this study assumed that not much variations in temperature and humidity. Hence, the model parameters for room temperature and relative humidity are set to 25° C and 30% respectively unless otherwise specified.

The viruses exhaled by an infected person contaminates the surroundings and that causes the virus spreading to nearby humans. In reality, the exhaled virus particles will have an initial velocity when they are ejected from the mouth or nose which depends on the flow velocity of the exhaled air. The flow velocity of exhaled air may vary from person to person depends on many parameters such as lungs capacity, eject during talking or in tidal breathing, physiological characteristics of airway, mouth and nose etc. This makes the system complex and hence this computer model assumes that initial velocity of exhaled particle is zero.

## Results

### Graph shows the distribution of virus around the infected person who is standing in the queue

**Figure 2:**
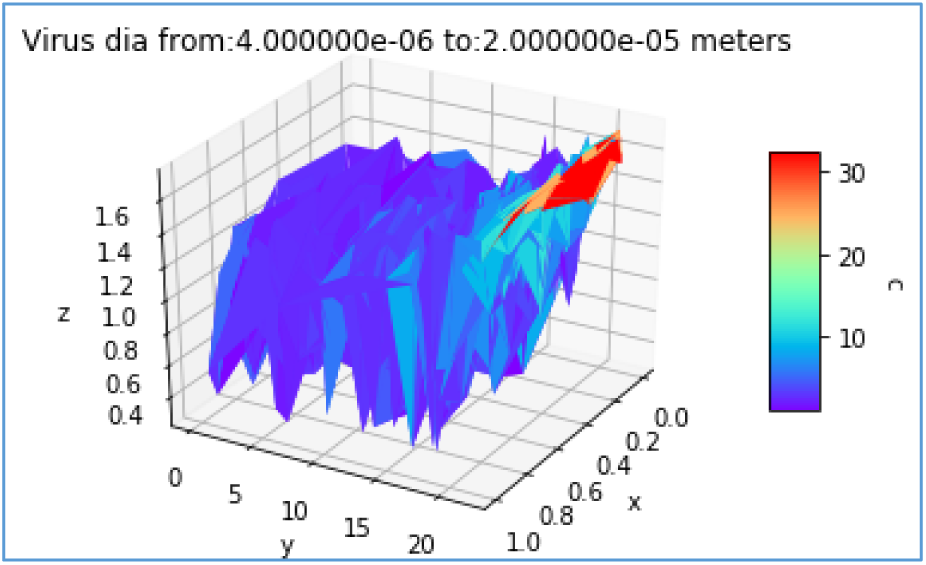
Distribution of virus particles (random diameter from 4uM to 20uM) Infected person moved from 1st to 20th position in the queue (y-axis); The virus particles settled slowly from the height of the release by the infected person (z-axis); The random distribution of particles across the queue (x-axis). The amount of virus particles in the space is shown in colour mapping. The area marked as Red contains the maximum number of particles. Please refer the colour map scale shown in the right side and the model parameters in the supporting information.

### Virus inhaled by a person behind the infected person in the queue

**Figure 3:**
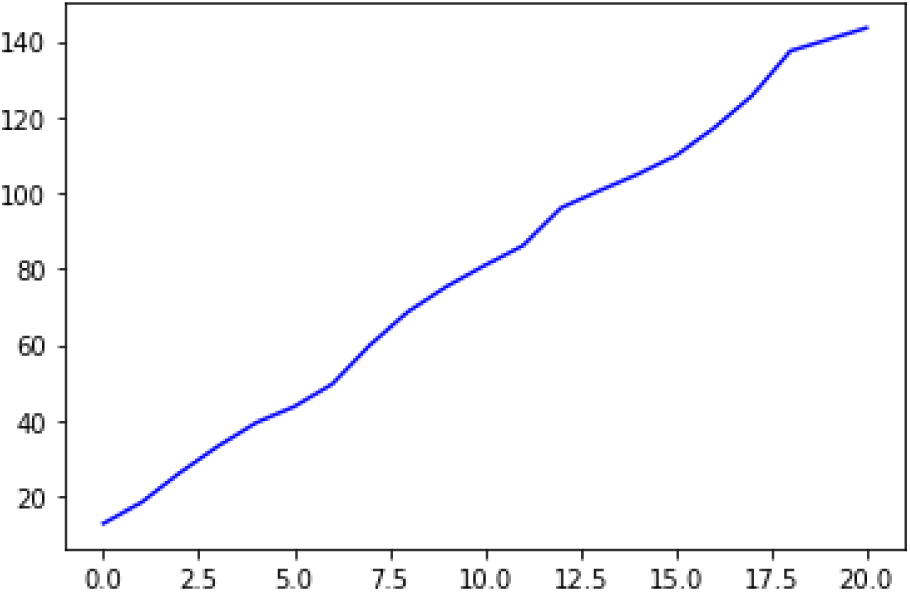
Cumulative inhaling of virus particles by the healthy person behind the infected person in the queue. The position of the healthy person in the queue is from 0.0 to 20.0 (x-axis); The total virus inhaled by the person is shown in the y-axis. Please refer model parameters in the supporting information.

### Relation of risk of infection with height of a person and the waiting time in the queue

The probability of getting infection to the person just behind the symptomatic or asymptomatic Covid-19 patient will increase as the waiting time in the queue. Waiting time is defined as the time taken to move forward one step in the queue. The probability of spreading Corona virus when the waiting time is 150 seconds is almost 8 times more than that of 30 seconds waiting time in the queue as shown in figure 4.

**Figure 4:**
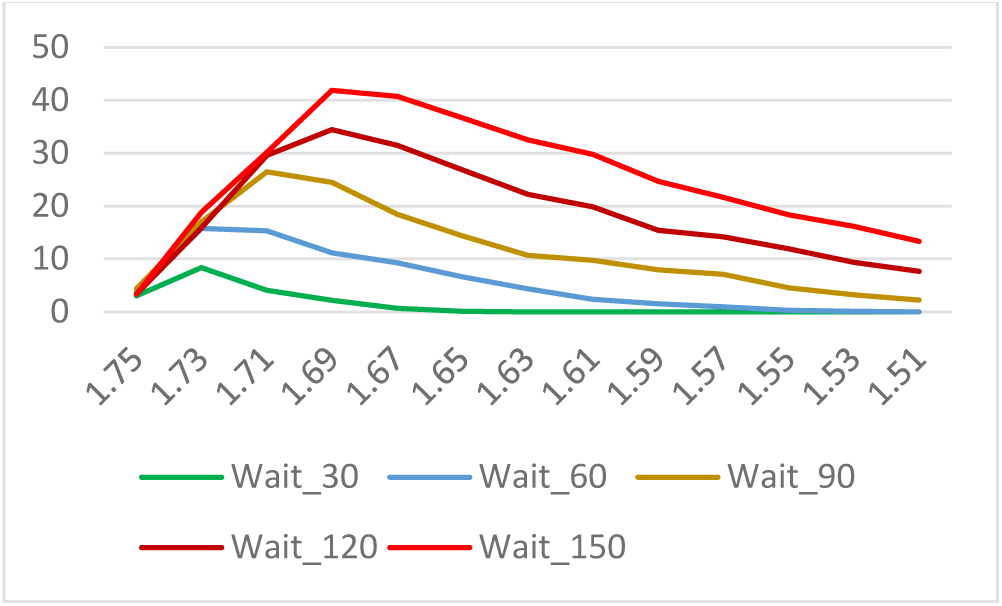
The virus consumption by the person behind the infected person in the queue; The height of the person is in the X-axis, waiting time in the queue (30, 60, 90, 120, 150 seconds) in different colours; The number of viruses inhaled is shown in the Y-axis. Please refer the supporting information for model parameters.

### Relation of risk of infection with height and position of a healthy person in the queue

**Figure 5:**
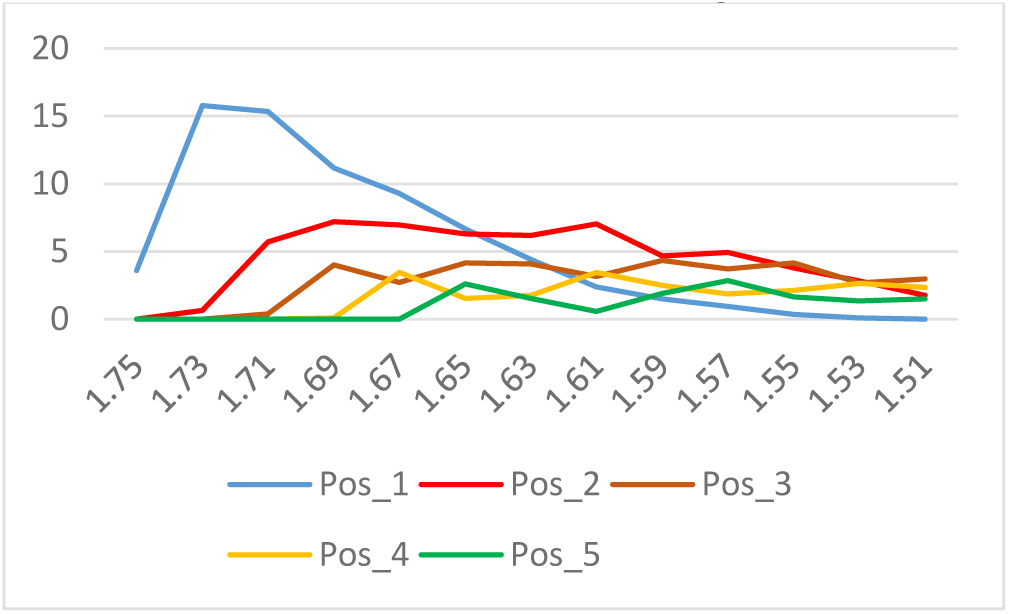
Vulnerability of the person behind the infected person in the queue; The height of the person is shown in the X-axis; The viruses inhaled by the person is shown in the Y-axis. The position of the person behind the infected person is shown in different colours. Position 1 is just behind the infected person, 2 means one another person is in between the subject and the infected person and so on. Please refer the model parameters in the supporting information. Waiting time: 60 seconds

**Figure 6:**
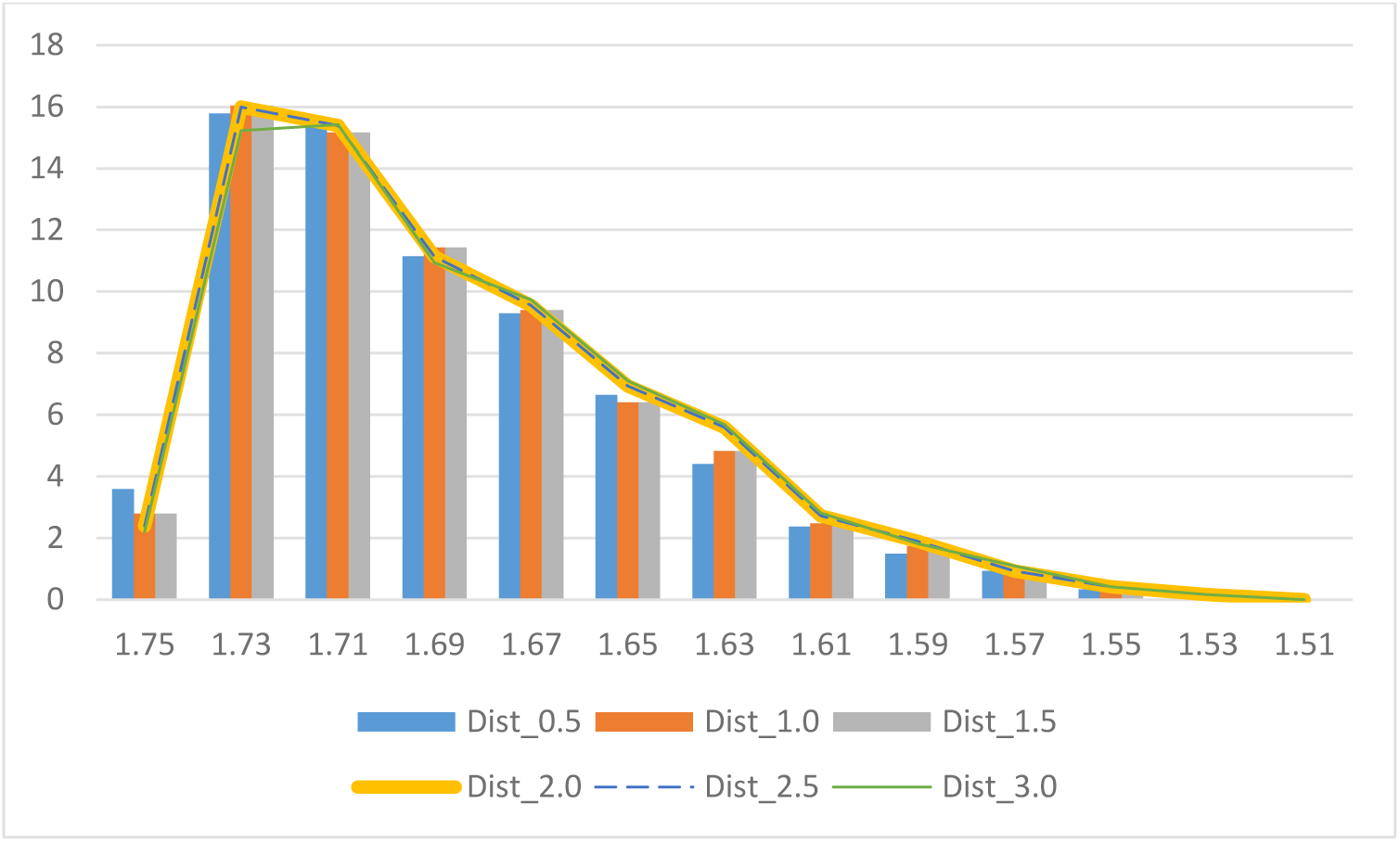
Effect on distance between people in the queue and their height. The number of virus particles inhaled by the healthy person shown in y-axis and the height of the person in x-axis; the distance between people in the queue is from 0.5 meter to 3 meters. The number of viruses inhaled by the person is irrespective of distances between people. For the clarity, the graph is shown in the combo of bar chart and line chart with different colour and width. The walking speed of the person in the queue is considered as 3.5 Km/hour even though the normal speed of a human is between 4.5 Km/Hour to 5.0 Km/hour. The small difference in the number of virus particles inhaled by a healthy person with the respect to the distance between people is due the delay in reaching to the next position. During this delay, due to the free fall, some of the virus particles can entry or exit from a particular air column.

## Discussion

Some of the present articles describes about the possibility of transmission of Corona viruses even from asymptomatic subjects, which make the preventive system too complex to safeguard the human population from the Covid-19 pandemic. This study is concentrated in analysing the risk of affecting Corona virus when people are standing in a queue.

While writing this paper, the research are underway and the knowledge about the transmission of Corona virus is very limited. A computer model is made to overcome the practical difficulties to study and analyse the way of spreading of the new Corona virus when the infected person is in the queue.

The computer model simulates the generation of Corona viruses with a given diameter (or random diameters), exhale them by the infected person standing in the queue in every breath and the free fall of viruses by considering the drag force and time slices. The scattering factor and the consume factor are also incorporated in the computer model to make its outcome as close to the reality.

The virus distribution in the surrounding area of an infected is determined using this computer simulation. Even though the possibilities and the number of viruses that can excrete during the exhale of a Covid-19 person in not known but some studies confirmed that even asymptomatic person can exhale viruses during the tidal breathing in addition to the droplets that expels during the talk, cough or sneeze. [13]

The settling velocity of the aerosol or droplets are determined in the model based on Stoke’s law. The free fall velocity determines the time at which it is suspended in the air before settled down on the surface. The settling velocity is determined by the diameter of the droplets, the evaporation of the water contents in the enveloped Corona virus decides a major factor of the respiratory infection spread of Covid-19. The room temperature and relative humidity will also play the role in the Corona airborne transmission at least in a few meters.

The release and movement of virus particles are considered in every time slice and the distribution of particles in time and space are determined. As shown in figure 2, the 3D map shows the concentration of particles are maximum at the exhale point, near to the mouth and nose and reduce gradually from the release point. The model predicts that the maximum possibility of getting infection is the person just behind the infected person in the queue. As the queue progresses, the healthy person behind the Covid-19 patient move ahead and will always occupy the most contaminated area which is shown in Red and Orange in figure 2. This increases the possibility of inhaling the virus particles many fold compare to other occupants in the queue.

The potential risk of a healthy person behind an infected person in the queue is determined by running the computer model in time slices. The healthy person in the queue will inhale a fraction of total viruses in the contaminated area which is defined by the "consume factor” model parameter. The inhaled viruses will first binding to a host receptor and then fusing viral and host membranes. Over a period of time, inhaled viruses are accumulated in the person and susceptible to Covdi-19 when the amount of virus particles exceeds the infectious dose. The simulation records the quantity of inhaled viruses when the person is just behind the infected person and following him from first position to the last position in the queue. The accumulation of viruses in the healthy person is shown in the figure 3.

More time the people spend in the queue is more risky due to the fact that the amount of viruses exhaled by the infected person and the amount of viruses inhaled by the healthy people are cumulative. To investigate this hypothesis, the simulation is done by the computer model with various waiting time in the queue. The waiting time defined as the time in seconds take to move to the next step in the queue and the results are shown in figure 4. As the waiting time increases, the susceptibility will also increase to all height groups. With an infected person of height 1.75 meters, most susceptible person is with a height of 1.73 meters when the waiting time is 30 seconds while the person with a height of 1.69 meters is at the highest risk if the weighting time is 150 seconds.

The contaminated area is more infectious to a healthy person when his/her nose and mouth are come in close to the area. In other words, the probability of infection depends on the height of the patient as well as that of the healthy person in the queue. To understand the relation between the vulnerability of a person and the height, this computer simulation is repeated with various heights including the height of the Covid-19 patient who is standing in the queue. As expected and as shown in the figure 5, Blue line, if the healthy person is just behind the infected person, position 1, will inhale more viruses than any other person in the queue.

The person who is in the second position from the infected person will inhale more viruses only if the height is much less than that of the infected person in the queue (Figure 5, Red line). This is justified by the fact that virus particles are free falling and the contaminated area will fall further as the person in the second position occupy the position of the infected person.

The effect on the probability of spreading the corona virus among the people based on the distance between them in the queue is also studied by this computer model. The simulation has been carried out with various distances from 0.5 meter to 3 meters and obtained the respective number of viruses inhaled by the person in the queue. As shown in the figure 6, in all cases, irrespective of the distance between the people in the queue, the number of viruses inhaled by the healthy person remains the same. This is a surprising finding since we believe that increasing the physical space between people will reduce the chance of spreading illness. But in reality, irrespective of the distance between the people, when people move forward as the queue progresses the healthy person will possess the same position as that of the infected person hence the risk of infection remains the same. The small difference seen in the graph is due to the time taken by a person to reach next position in the queue and during this time some virus particles can reach or leave a particular section in the air column.

This computer simulation reveals the fact that the possibility of respiratory transmission of Corona virus is high at least in the considered model of a queue where symptomatic or asymptomatic Covid-19 patient is the part of it. The probability of high risk depends on the height of the infectious person and other people, position of the person and the waiting time in the queue. This fact forces us to think that the social distancing and personal hygiene alone are not sufficient to prevent Covid-19 but require to take other precautionary measures such as wearing facemasks with appropriate standards to prevent the respiratory transmission of Corona viruses.

## Conclusion

The developed computer model was used to predict the influence of several parameters on the droplets and environmental factors. A study has been conducted to understand the possibilities of spreading of Corona viruses among people standing in the queue in indoor controlled environment. The possibility of getting infection of a person in a queue depends on many factors; height of the infected person and occupants in the queue; position of the infected person in the queue; environmental factors such as room temperature and relative humidity. It was found that, the settling velocity of droplets decides the distribution of virus particles which in turn depends on the effective diameter of droplets. In this study, it is concluded that the people in the queue behind the infected person will be affected more and the severity depends on the height and the position of the person in the queue. Also observed that the people are more susceptible to Covid-19 when they spend more time in the queue if an infected person is standing in the queue. The distance between people in the queue is not having much effect on the probability of spreading of corona virus at least with the parameters used in this study.

Future research should further develop and confirm these initial findings by performing a detailed experimental study on the respiratory transmission of the new Human Corona Virus.

## Data Availability

It is included in the manuscript under the section “Supporting information”

## Funding Acknowledgements

The author received no financial support for the research, authorship, and/or publication of this article. This study has been conducted for the personal interest of the author and these activities emanates from a genuine interest to help the society by contributing the knowledge that may help to prevent the rapidly spreading Covid-19 pandemic across the globe.

## Supporting information

Default Model parameters

| Parameter | Description | Default value | Dimension |
| --- | --- | --- | --- |
| $V_{\text{exhale}}$ | No. of virus per exhale | 5 | Nil |
| $\text{Breath}_{\text{rate}}$ | Breath rate of human | 12 | Per minute |
| $Q_{\text{Wait}}$ | Waiting time in the queue | 60 | s |
| $Q_{\text{Length}}$ | Queue Length | 20 | m |
| $Q_{\text{Dist}}$ | Distance between people in Queue | 1 | m |
| $\text{Walk}_{\text{speed}}$ | Walking speed of a person | 4.5 | Km/hour |
| $\text{Patient}_{\text{Ht}}$ | Height of patient | 1.75 | m |
| $\text{Healthy}_{\text{Ht}}$ | Height of healthy person | 1.75 | m |
| $\text{Healthy}_{\text{Pos}}$ | Position of healthy person from patient in the queue | 1 | Nil |
| $\text{Cons}_{\text{fact}}$ | Consume factor | 0.1 | Nil |
| $\text{Scatter}_{\text{fact}}$ | Scatter factor per 0.1 meter | 0.1 | $\text{m}^{-1}$ |
| $\text{VirusDia}_{\text{Start}}$ | Start Virus dia for random dia | 4.0e-06 | m |
| $\text{VirusDia}_{\text{End}}$ | End Virus dia for random dia | 20e-06 | m |
| $V_{\text{CrossWind}}$ | Crosswind velocity | 0.0 | $\text{ms}^{-1}$ |
| $A_{\text{CossWind}}$ | Crosswind angle from ground | 0.0 | degree |
| $V_{\text{FrontWind}}$ | Front wind velocity | 0.0 | $\text{ms}^{-1}$ |
| $A_{\text{FrontWind}}$ | Front wind angle | 0.0 | degree |
| $\text{Room}_{\text{Temp}}$ | Room temperature | 25 | $^{\circ}\text{C}$ |
| $\text{Room}_{\text{RH}}$ | Room Relative humidity | 30 | % |

Model parameters of Figure 2:

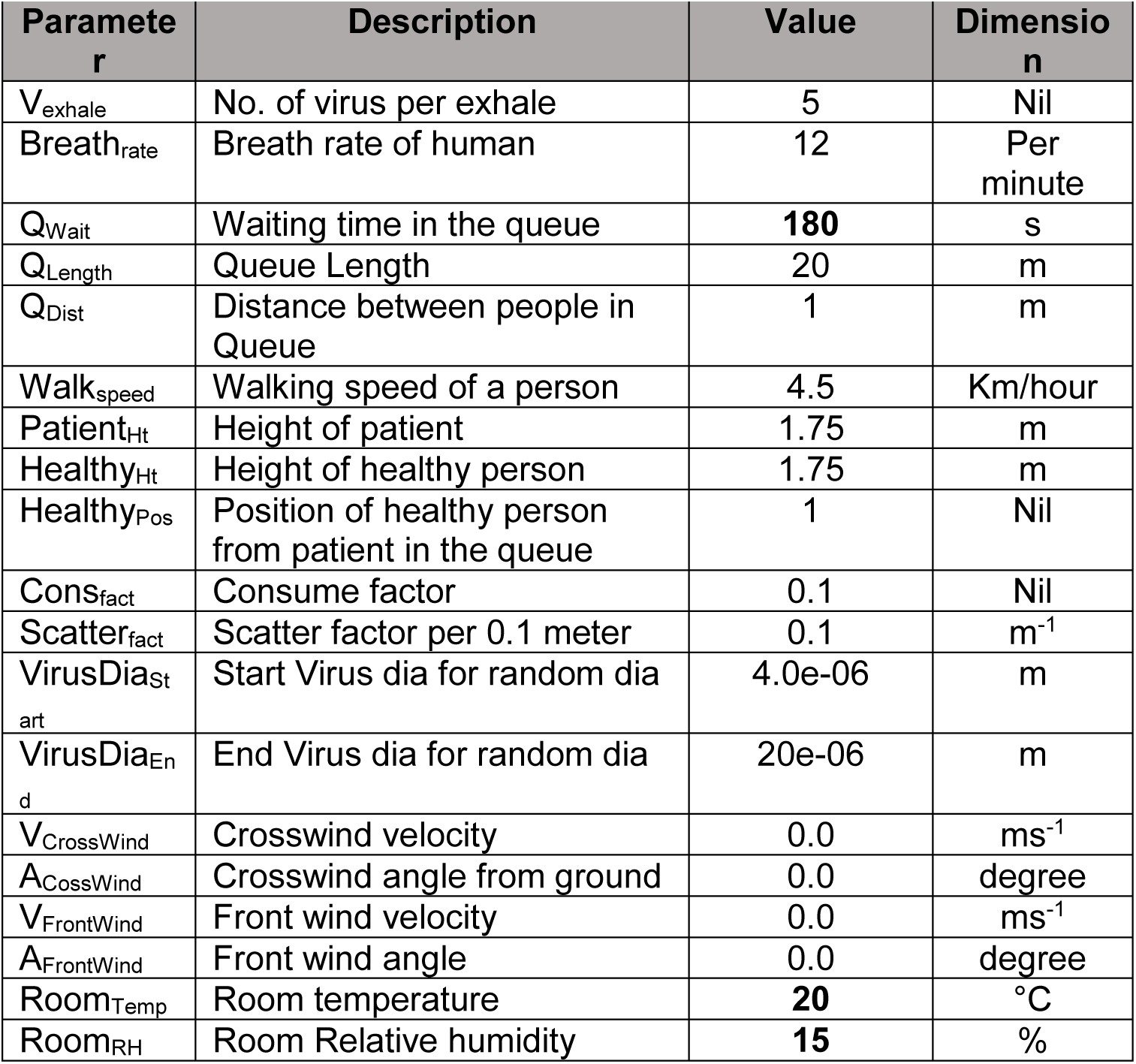

Model parameters of Figure 3:

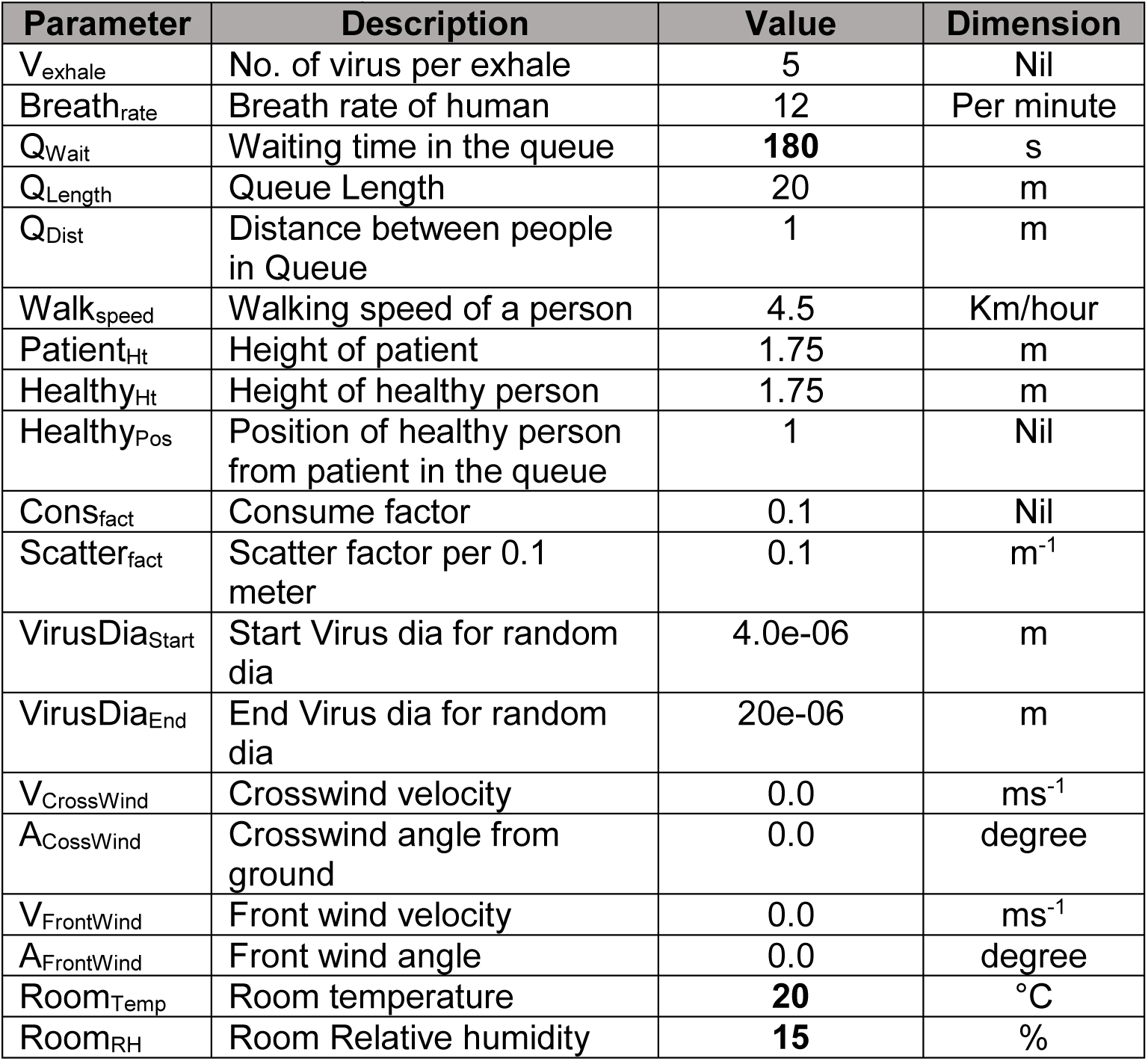

Model parameters of Figure 4:

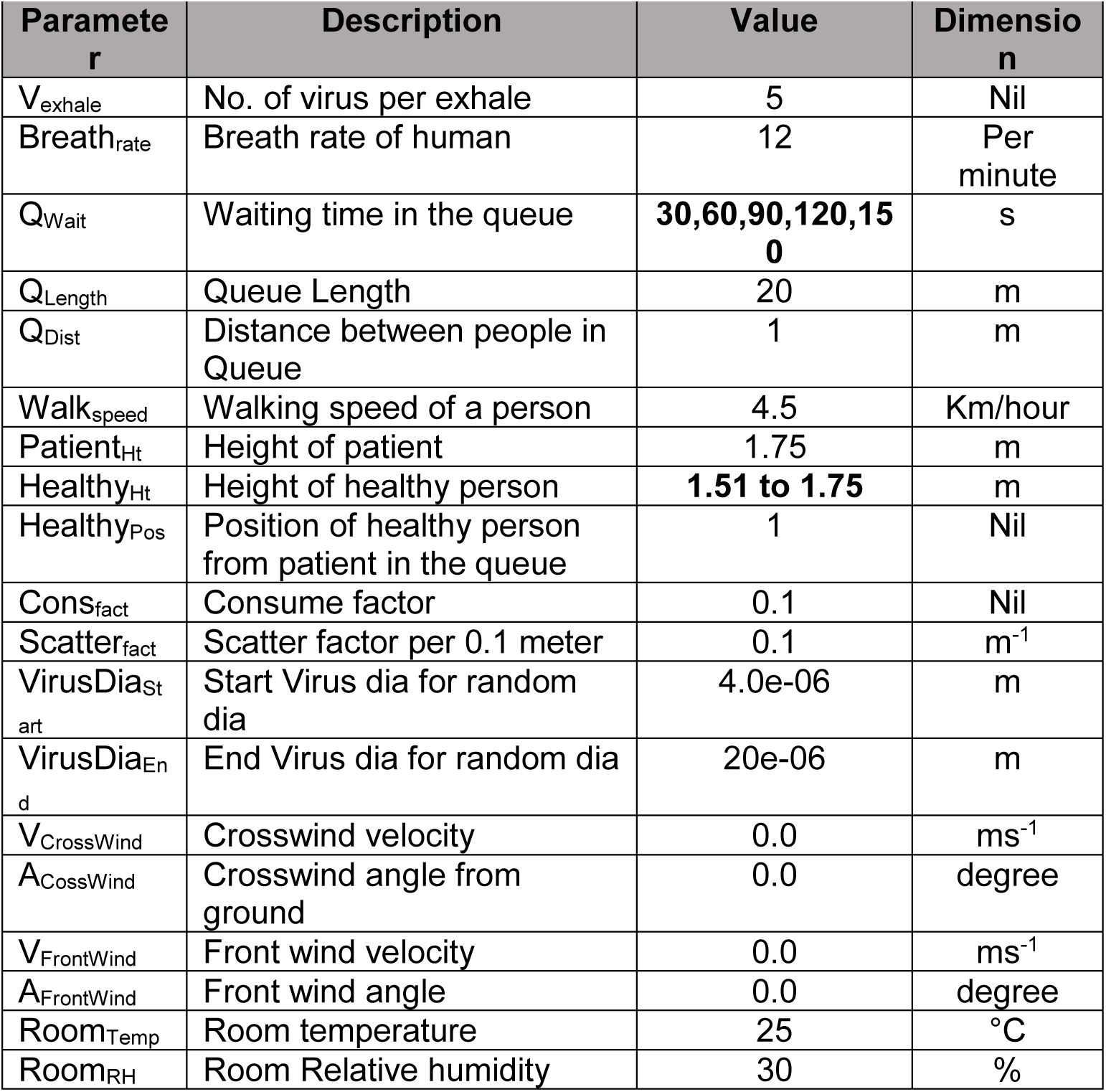

Model parameters of Figure 5:

| Parameter | Description | Value | Dimension |
| --- | --- | --- | --- |
| $V_{\text{exhale}}$ | No. of virus per exhale | 5 | Nil |
| $\text{Breath}_{\text{rate}}$ | Breath rate of human | 12 | Per minute |
| $Q_{\text{Wait}}$ | Waiting time in the queue | 60 | s |
| $Q_{\text{Length}}$ | Queue Length | 10 | m |
| $Q_{\text{Dist}}$ | Distance between people in Queue | 1 | m |
| $\text{Walk}_{\text{speed}}$ | Walking speed of a person | 4.5 | Km/hour |
| $\text{Patient}_{\text{Ht}}$ | Height of patient | 1.75 | m |
| $\text{Healthy}_{\text{Ht}}$ | Height of healthy person | **1.51 to 1.75** | m |
| $\text{Healthy}_{\text{Pos}}$ | Position of healthy person from patient in the queue | **1, 2, 3, 4, 5** | Nil |
| $\text{Cons}_{\text{fact}}$ | Consume factor | 0.1 | Nil |
| $\text{Scatter}_{\text{fact}}$ | Scatter factor per 0.1 meter | 0.1 | $\text{m}^{-1}$ |
| $\text{VirusDia}_{\text{Start}}$ | Start Virus dia for random dia | 4.0e-06 | m |
| $\text{VirusDia}_{\text{End}}$ | End Virus dia for random dia | 20e-06 | m |
| $V_{\text{CrossWind}}$ | Crosswind velocity | 0.0 | $\text{ms}^{-1}$ |
| $A_{\text{CossWind}}$ | Crosswind angle from ground | 0.0 | degree |
| $V_{\text{FrontWind}}$ | Front wind velocity | 0.0 | $\text{ms}^{-1}$ |
| $A_{\text{FrontWind}}$ | Front wind angle | 0.0 | degree |
| $\text{Room}_{\text{Temp}}$ | Room temperature | 25 | $^{\circ}\text{C}$ |
| $\text{Room}_{\text{RH}}$ | Room Relative humidity | 30 | % |

Model parameters of Figure 6:

| Parameter | Description | Default value | Dimension |
| --- | --- | --- | --- |
| $V_{\text{exhale}}$ | No. of virus per exhale | 5 | Nil |
| $\text{Breath}_{\text{rate}}$ | Breath rate of human | 12 | Per minute |
| $Q_{\text{Wait}}$ | Waiting time in the queue | 60 | s |
| $Q_{\text{Length}}$ | Queue Length | 10 | m |
| $Q_{\text{Dist}}$ | Distance between people in Queue | **0.5, 1.0, 1.5, 2.0, 2.5, 3.0** | m |
| $\text{Walk}_{\text{speed}}$ | Walking speed of a person | **3.5** | Km/hour |
| $\text{Patient}_{\text{Ht}}$ | Height of patient | 1.75 | m |
| $\text{Healthy}_{\text{Ht}}$ | Height of healthy person | **1.51 to 1.75** | m |
| $\text{Healthy}_{\text{Pos}}$ | Position of healthy person from patient in the queue | 1 | Nil |
| $\text{Cons}_{\text{fact}}$ | Consume factor | 0.1 | Nil |
| $\text{Scatter}_{\text{fact}}$ | Scatter factor per 0.1 meter | 0.1 | $\text{m}^{-1}$ |
| $\text{VirusDia}_{\text{Start}}$ | Start Virus dia for random dia | 4.0e-06 | m |
| $\text{VirusDia}_{\text{End}}$ | End Virus dia for random dia | 20e-06 | m |
| $V_{\text{CrossWind}}$ | Crosswind velocity | 0.0 | $\text{ms}^{-1}$ |
| $A_{\text{CossWind}}$ | Crosswind angle from ground | 0.0 | degree |
| $V_{\text{FrontWind}}$ | Front wind velocity | 0.0 | $\text{ms}^{-1}$ |
| $A_{\text{FrontWind}}$ | Front wind angle | 0.0 | degree |
| $\text{Room}_{\text{Temp}}$ | Room temperature | 25 | $^{\circ}\text{C}$ |
| $\text{Room}_{\text{RH}}$ | Room Relative humidity | 30 | % |

## Notes

### Competing Interest Statement

The authors have declared no competing interest.

### Author Declarations

This computer simulation study does not involve any clinical trial

